# Neonatal Respiratory Morbidity among Late Preterm Births by Planned Mode of Delivery and Gestational Age

**DOI:** 10.1101/2024.01.04.23299873

**Authors:** Mark A. Clapp, Siguo Li, Jessica L. Cohen, Cynthia Gyamfi-Bannerman, Amy B. Knudsen, Scott A Lorch, Tanayott Thaweethai, Jason D. Wright, Anjali J. Kaimal, Alexander Melamed

## Abstract

**Objective:** To estimate the effect of late preterm antenatal steroids on the absolute risk of respiratory morbidity among subgroups of patients based on the planned mode of delivery and gestational age at presentation.

**Methods:** This was a secondary analysis of the Antenatal Late Preterm Steroid (ALPS) Trial, a multicenter trial originally conducted within the NICHD’s MFMU network of individuals with singleton gestations and without pre-existing diabetes who were at high risk for late preterm delivery (34-36 weeks of gestation). We fit binomial regression models to estimate the absolute risk of respiratory morbidity, with and without steroid administration, by gestational age and planned mode of delivery at the time of presentation. We assumed a homogenous effect of steroids on the log-odds scale, as was reported in the ALPS trial. The primary outcome was neonatal respiratory morbidity, as defined in the ALPS Trial

**Results:** The analysis included 2,825 patients at risk for late preterm birth. The risk of respiratory morbidity varied significantly by planned mode of delivery (adjusted risk ratio (aRR) 1.90 (95% confidence interval (CI) 1.55, 2.33) for cesarean compared to vaginal delivery) and day of gestation at presentation (aRR 0.92 (95% CI 0.90, 0.94)). For those planning a cesarean delivery and presenting in the 34th week of gestation, the risk of neonatal respiratory morbidity was 39.4% (95% CI 30.8, 47.9%) without steroids and 32.0% (95% CI 24.6, 39.4%) with steroids. In contrast, for patients presenting in the 36th week and planning a vaginal delivery, the risk of neonatal respiratory morbidity was 6.9% (95% CI 5.6, 8.6%) without steroids and 5.6% (95% 4.2, 7.0%) with steroids.

**Conclusion:** The absolute risk of neonatal respiratory morbidity among patients at risk for late preterm delivery varies considerably by gestational age at presentation and planned mode of delivery. As only communicating the relative risk reduction of antenatal steroids for respiratory morbidity may lead to an inaccurate perception of benefit, individualized estimates of absolute risk expected with and without treatment may inform shared decision-making.

## Introduction

Neonatal respiratory morbidity, including respiratory distress syndrome and transient tachypnea of the newborn, is a common complication of prematurity.^1–3^ Multiple patient factors, including gestational, mode of delivery, and fetal sex, influence the risk of respiratory morbidity.^2–7^ There is a broad and longstanding consensus that antenatal steroids decrease the risk of neonatal respiratory morbidity for neonates born before 34 weeks of gestation.^8,9^ More recently, the Antenatal Late Preterm Steroid (ALPS) Trial demonstrated that this benefit extends to fetuses at risk of delivery in the late preterm period (34-36 weeks of gestation): neonates exposed antenatally to betamethasone in the late preterm period had a 20% risk reduction of the composite respiratory outcome, without statistically conclusive evidence of treatment heterogeneity among pre-specified subgroups.^10^

The Society for Maternal-Fetal Medicine recommends that clinicians and patients engage in shared decision-making around steroid administration in the late preterm period.^11^ However, the baseline risk of respiratory morbidity differs substantially across the late preterm period, making summary measures of absolute risk, as reported in the original trial, less informative for individualized counseling.^2^ Furthermore, communicating only the relative risk may lead to an inaccurate perception of the benefit of antenatal steroids, especially for those with a small absolute risk.^12^ Thus, our objective in this secondary analysis of data from the ALPS Trial was to estimate the absolute risk of respiratory morbidity with and without steroid administration, based on patients’ planned mode of delivery and gestational age at presentation.^10^

## Methods

This is a secondary analysis of the Antenatal Late Preterm Steroid (ALPS) Trial conducted within 17 university-based hospitals within the Maternal-Fetal Medicine Units (MFMU).^10^ In the trial, patients with a singleton pregnancy between 34 weeks 0 days and 36 weeks 5 days of gestation and with a high chance of delivery in the late preterm period were enrolled and randomized to receive an antenatal course of betamethasone or placebo. The primary outcome was a composite of respiratory morbidity within 72 hours after delivery and included the following: high-flow nasal cannula or continuous positive airway pressure (CPAP) for at least 2 hours; supplemental oxygen use for at least 4 hours; extracorporeal membrane oxygenation (ECMO); and/or mechanical ventilation. We also examined the secondary outcome of severe respiratory complications, as defined in the ALPS Trial as “a composite outcome of CPAP or high-flow nasal cannula for at least 12 continuous hours, supplemental oxygen with a fraction of inspired oxygen of at least 0.30 for at least at least 24 continuous hours, ECMO or mechanical ventilation, stillbirth, or neonatal death within 72 hours after delivery.”^10^ The ALPS trial randomized 2,831 patients to betamethasone (n=1,429) or placebo (n=1,402).^10^ This analysis included patients randomized at or after 34 weeks of gestation who were not lost to follow-up.

We examined the risk of neonatal respiratory morbidity among two patient factors strongly associated with the risk of respiratory morbidity: mode of delivery and gestational age.^2,3,5^ To generate results that could guide clinical decision-making and inform counseling, we considered *planned* mode of delivery (not actual mode) and gestational age *at presentation* (not delivery), information available to clinicians and patients when deciding about antenatal steroid administration. We defined the planned mode of delivery as “cesarean” if 1) a cesarean delivery was scheduled in the late preterm period at the time of presentation or 2) if no delivery was scheduled at the time of presentation but a cesarean section was performed at any gestational age for the following indications: placenta accreta, placenta previa, fetal anomaly, prior uterine surgery, or prior cesarean delivery. This definition was chosen to capture patients who were unlikely to attempt a trial of labor. The prevalence of these factors was compared between the two groups to examine for potential group imbalances in the original trial.

We constructed a binomial regression model that included the treatment assignment (placebo or betamethasone), planned mode of delivery, and gestational age in days and used a log link to report risk ratios. Because the prespecified subgroup analysis in the original trial did not detect signs of treatment effect heterogeneity, the model assumed a constant effect size (on the log-odds scale) of the intervention across all groups. We used this regression model to estimate the absolute risks of respiratory morbidity in both the treated and untreated groups. The predicted (or estimated) absolute risks of this model were presented graphically by planned mode of delivery and across days of gestation; for comparison, we also plotted the observed absolute risks of the outcome with a corresponding 95% confidence interval by planned mode of delivery and weeks of gestation. To examine the model’s discrimination and calibration, we used k-fold cross-validation (k=10) to calculate the area under the receiver operating characteristic (ROC) curves (AUC) and generate a calibration plot for both outcomes.

Additional steps were performed to further facilitate counseling around the absolute risk of respiratory morbidity and expected outcome by mode of delivery and gestational age. First, the risks of the primary and secondary outcomes were estimated using the same logistic regression model, though gestational age in weeks was used instead of gestational age in days; this allowed for the output to be summarized in a compact, interpretable table. Second, we built an online reference tool to display the estimated risk of the primary and secondary outcome using a patient’s precise gestational age in days at presentation and their planned mode of delivery, which is available at https://mghinferencelab.shinyapps.io/alps_web_calc/.

Data was obtained with permission from the NICHD Data and Specimen Hub (DASH).^13^ The XXX Institution Review Board exempted this study. All analyses used Stata (Stata Corp, College Station, TX). P-values <0.05 were considered statistically significant. The STROBE reporting guidelines were followed.

## Results

Of the original 2,831 participants, we excluded 2 patients randomized before 34 weeks of gestation and 4 patients lost to follow-up (2.1%). eTable 1 in the Supplement shows the sample sizes of the various strata used in the analysis. As expected, there were no differences in distributions of the planned mode of delivery or gestational age at randomization between the two groups (eTable 2 in the Supplement).

Figure 1 shows the predicted absolute risks of respiratory morbidity among patients treated with steroids and those who did not by the day of gestation and the planned mode of delivery at the time of presentation. The observed rates (not those predicted by the model) by week of gestation are shown on the same graph for comparison and included in eTable 3 in the Supplement. The risk of respiratory morbidity varied significantly across planned modes of delivery (adjusted risk ratio (aRR) 1.90 (95% confidence interval (CI) 1.55, 2.33) for cesarean compared to vaginal delivery) and day of gestation at presentation (aRR 0.92 (95% CI 0.90, 0.94)). The unadjusted and adjusted model outputs are shown in eTable 4 in the Supplement. The 10-fold cross-validated AUC was 0.669 (95% CI 0.639, 0.698). The ROC curve and calibration plot is shown in eFigure 1 and eFigure 2 in the Supplement.

**Figure 1:**
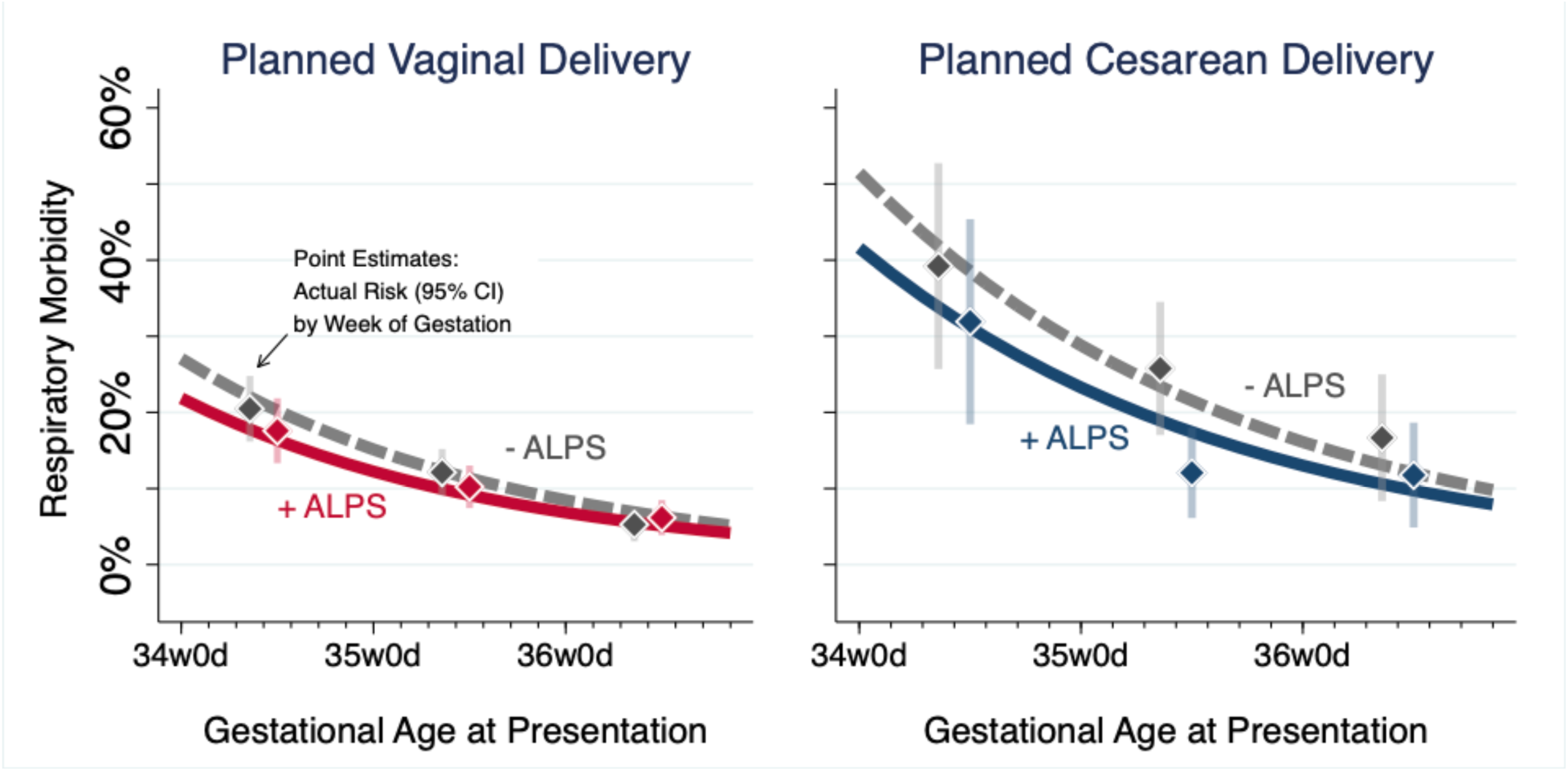
Risk of respiratory morbidity among individuals exposed and not exposed to antenatal steroids by gestational age and planned mode of delivery The lines represent the estimated rates of respiratory morbidity among those exposed (+ ALPS) and not exposed (-ALPS) that were derived from a logistic regression model that included gestation age at presentation (in days), planned mode of delivery (vaginal vs. cesarean), and treatment group. For comparison, the observed rates (i.e., “actual risk”) with their corresponding 95% confidence interval (CI) by week (not day) of gestation are also plotted; of note, their position on the x-axis does not correspond to a specific day within the week of gestation.

The predicted absolute risks with and without treatment are summarized by week of gestation in Table 1. At baseline (i.e., among those treated with the placebo), neonates born to individuals planning a cesarean delivery have a 36.7% (95% CI 29.8, 43.7%) risk of respiratory morbidity when presenting in the 34^th^ week of gestation; this risk decreases to 31.4% (95% CI 24.5, 37.9%) with antenatal steroids. By contrast, neonates who are born via a planned vaginal delivery after presenting in the 36^th^ week of gestation have a 6.8% (95% CI 5.1, 8.5%) risk of respiratory morbidity without steroids and a 5.4% (95% CI 4.0, 6.9%) risk with steroid treatment.

**Table 1:** Estimated risk of respiratory morbidity among neonates not exposed and exposed to antenatal steroids, stratified by week of gestation and planned mode of delivery

| Week of Gestation at Presentation | Planned Vaginal Delivery |  | Planned Cesarean Delivery |  |
| --- | --- | --- | --- | --- |
|  | No ALPS<br>% (95% CI) | ALPS<br>% (95% CI) | No ALPS<br>% (95% CI) | ALPS<br>% (95% CI) |
| 34 weeks | 20.9 (17.5-24.4) | 17.0 (13.9-20.2) | 39.4 (30.8, 47.9) | 32.0 (24.6-39.4) |
| 35 weeks | 11.8 (9.6-14.1) | 9.6 (7.7-11.5) | 22.3 (17.3-27.3) | 18.1 (14.0-22.3) |
| 36 weeks | 6.9 (5.2-8.6) | 5.6 (4.2-7.0) | 12.9 (9.3-16.6) | 10.6 (7.5-13.) |

ALPS, antenatal late preterm steroids; CI, confidence interval.

Figure 2 shows the predicted absolute risks of severe respiratory complications among those who received and did not receive antenatal steroids by day of gestational age and planned mode of delivery. The observed risks are shown on the same graphs by week of gestation and are included in eTable 5 in the Supplement. Planned mode of delivery (aRR 2.16 (95% CI 1.72, 2.72) for cesarean vs. vaginal delivery) and gestational age (aRR 0.92 (95% CI 0.90, 0.94)) were significantly associated with the risk of severe respiratory complications. The unadjusted and adjusted model outputs are shown in eTable 6 in the Supplement. The 10-fold cross-validated AUC was 0.679 (95% CI 0.651, 0.717); the ROC curve and calibration plots are shown in eFigure 3 and eFigure 4 in the Supplement. These findings are summarized by week of gestation in Table 2 which tabulates the absolute risk of severe respiratory complication, with and without treatment, by planned mode of delivery and week of gestational age at presentation.

**Figure 2:**
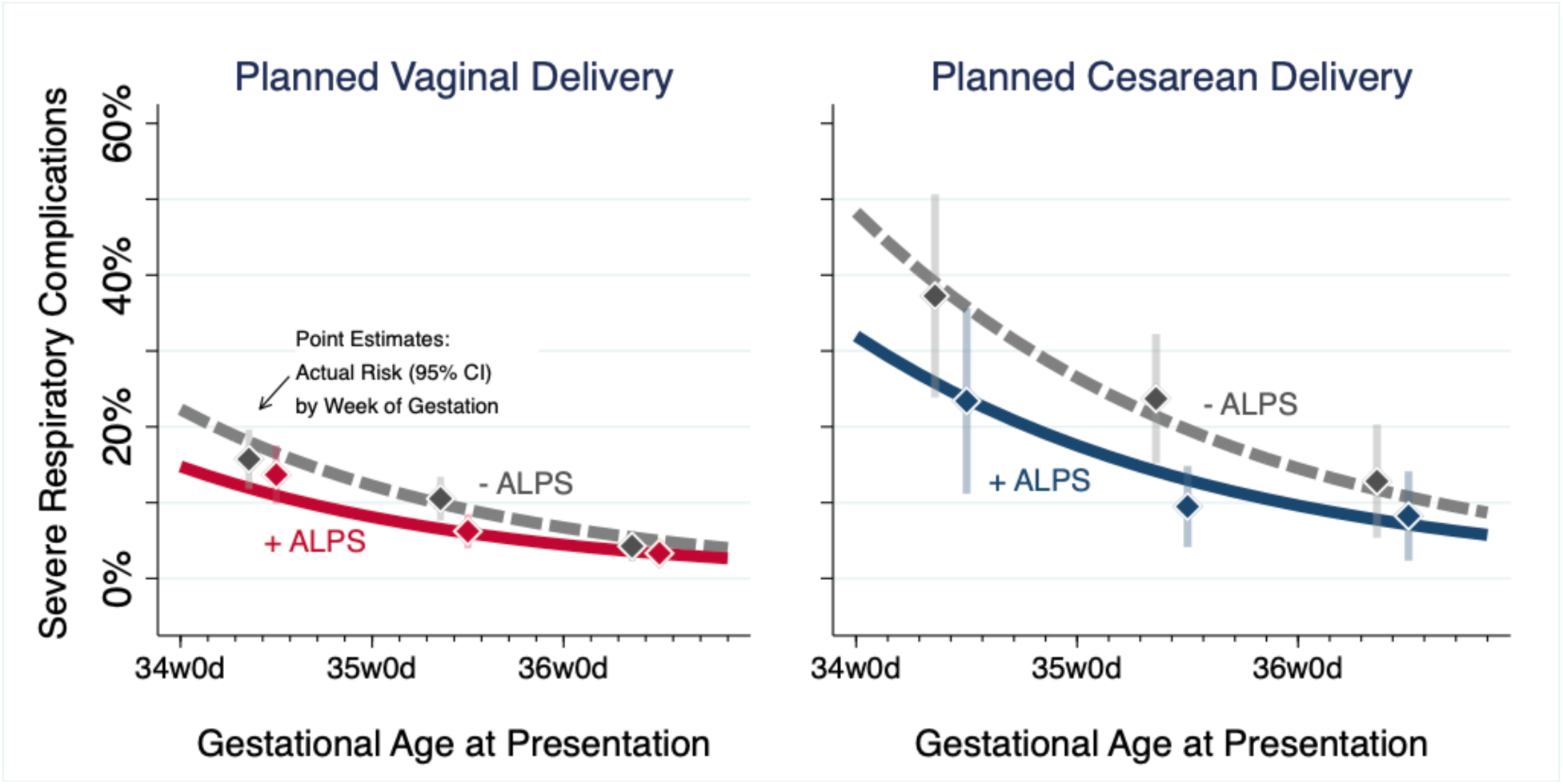
Risk of severe respiratory complications among individuals exposed and not exposed to antenatal steroids by gestational age and planned mode of delivery The lines represent the estimated rates of severe respiratory complications among those exposed (+ ALPS) and not exposed (-ALPS) that were derived from a logistic regression model that included gestation age at presentation (in days), planned mode of delivery (vaginal vs. cesarean), and treatment group. For comparison, the observed rates (i.e., “actual risk”) with their corresponding 95% confidence interval (CI) by week (not day) of gestation are also plotted; of note, their position on the x-axis does not correspond to a specific day within the week of gestation.

**Table 2:** Estimated risk of severe respiratory complications among neonates not exposed and exposed to antenatal steroids, stratified by week of gestation and planned mode of delivery

| Week of Gestation at Presentation | Planned Vaginal Delivery |  | Planned Cesarean Delivery |  |
| --- | --- | --- | --- | --- |
|  | No ALPS<br>% (95% CI) | ALPS<br>% (95% CI) | No ALPS<br>% (95% CI) | ALPS<br>% (95% CI) |
| 34 weeks | 17.3 (14.1-20.1) | 11.6 (9.0-14.2) | 37.0 (28.1-45.9) | 24.8 (18.1-31.4) |
| 35 weeks | 9.4 (7.6-11.8) | 6.5 (5.0-8.1) | 20.8 (15.6-25.9) | 13.9 (10.2-17.6) |
| 36 weeks | 5.0 (3.5-6.4) | 3.3 (2.3-4.4) | 10.6 (7.1-14.1) | 7.1 (4.7-9.5) |

ALPS, antenatal late preterm steroids; CI, confidence interval.

The results of these two models are available as an interactive calculator available at https://mghinferencelab.shinyapps.io/alps_web_calc/. This tool displays the estimated absolute risk of respiratory morbidity (and associated 95% CIs) with and without treatment as a function of day of gestation at presentation and planned mode of delivery (example shown in Figure 3).

**Figure 3:**
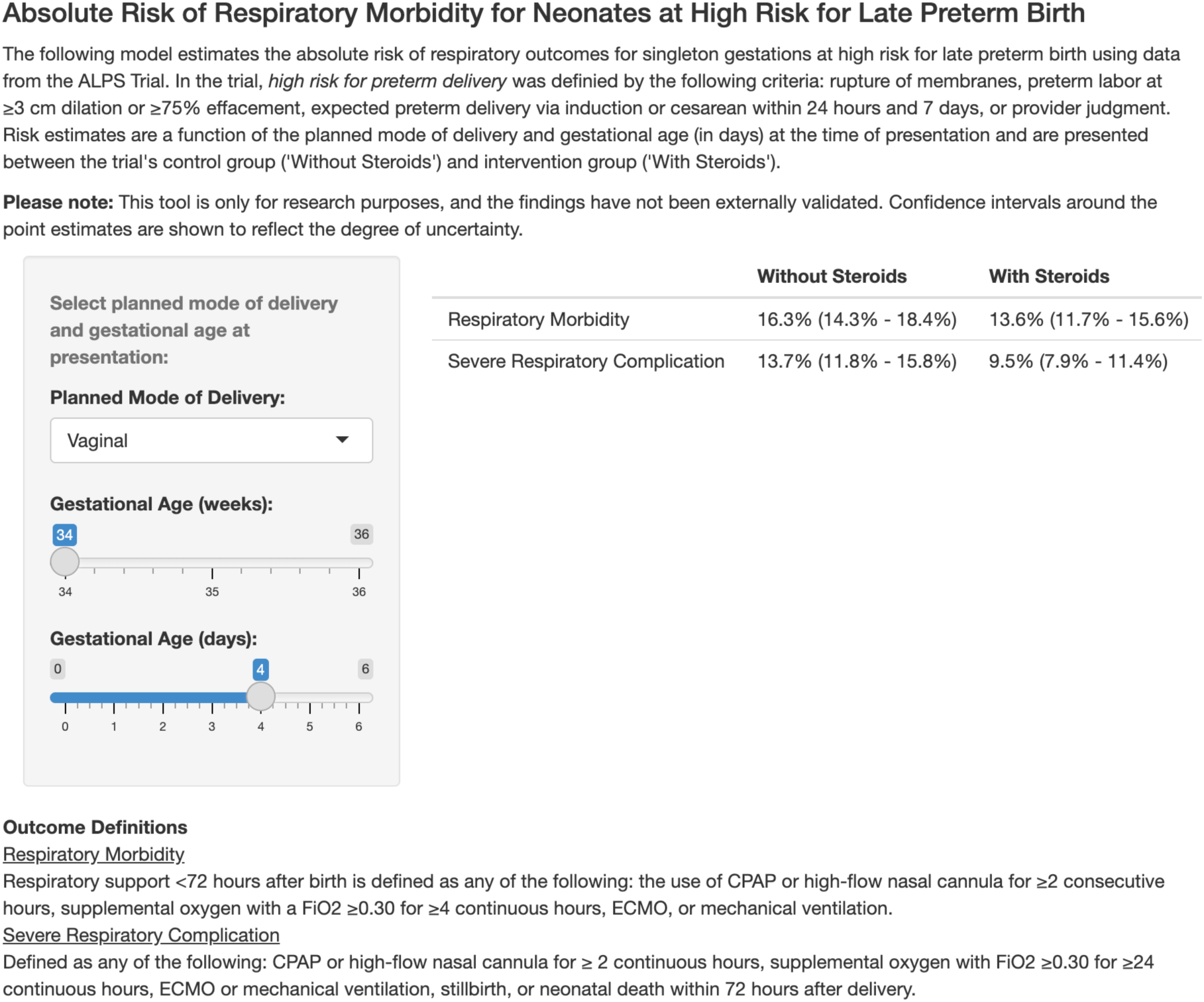
Image of online calculator to estimate the absolute risks of respiratory morbidity by planned mode of delivery and gestational age with and without antenatal steroids

## Discussion

In this secondary analysis of the ALPS Trial, we demonstrated that the absolute risks of neonatal respiratory morbidity vary significantly by gestational age and planned mode of delivery. Notably, infants born via a planned cesarean delivery are at a much higher risk of respiratory morbidity than those having a vaginal delivery, especially at earlier gestational ages (approximately 37% for planned cesarean delivery at 34 weeks vs. 7% for planned vaginal delivery at 36 weeks). While these relationships were previously known, we quantified this risk in the late preterm period using prospectively collected data as part of the ALPS Trial. Similarly, we demonstrated the challenge of predicting these outcomes for patients at the time of counseling and decision-making for antenatal steroids, even in the context of a clinical trial that used a strict definition of “high risk for preterm delivery,” as demonstrated by the ROC curves; to convey the level of uncertainty around the estimates, all data are presented with 95% confidence intervals. This information can inform shared decision-making about late preterm steroids and allow clinicians to estimate the baseline risk of neonatal respiratory morbidity more accurately. This absolute baseline risk of respiratory in the unexposed group can then be used to interpret the expected relative risk reduction that was demonstrated uniformly across all enrollees in the trial.

These findings must be interpreted in the context of the original ALPS Trial protocol and the trial enrollees. The ALPS Trial only enrolled patients who were at high risk of a late preterm delivery as determined by predefined clinical factors (rupture of membranes, preterm labor at ≥3 cm dilation or ≥75% effacement, or expected preterm delivery via induction or cesarean within 24 hours and 7 days) or provider judgment who had not previously received antenatal corticosteroids or had pregestational diabetes.^10^ As a result, nearly 40% of enrollees delivered before receiving the second dose, and the median time from presentation to delivery was approximately 30 hours.^10^ The absolute risks reported from this trial are most relevant to patients presenting with a similar risk of preterm delivery. For example, the benefit of antenatal steroids may be higher than reported in the trial for patients who are more likely to deliver after receiving a full course; in contrast, the benefit may be less if more patients received antenatal steroids but went on to delivery at later gestations or full term (i.e., a lower risk of preterm delivery than the trial). Studies of late preterm antenatal steroids outside of the context of a clinical trial are needed to better estimate the absolute risks of respiratory morbidity and the benefit of the intervention.

The original trial did include subgroup comparisons for several factors, including planned cesarean delivery and gestational age, though gestational age was analyzed as 34-35 vs. 36 weeks of gestation. The study reported no significant effects for the interaction terms for the primary outcome when each factor was analyzed individually; however, the trial was not powered to detect differences in the primary outcome within subgroups. The possibility of a differential impact of betamethasone in actual practice (i.e., treatment effect heterogeneity) remains plausible, especially considering the differences in the number of days between first dose and delivery and the number of patients who received a full course of betamethasone before delivery by planned mode of delivery and gestational age at presentation. Investigations of treatment effect heterogeneity should be a focus of future research to further individualize counseling on risks and benefits.

This study focused on counseling regarding the individualized benefits of antenatal steroids among two common and known clinical factors across which the baseline risk of respiratory morbidity varies widely. However, other factors may inform clinical decision-making, especially the known and theoretical intervention risks. Hypoglycemia, which was more likely in the treatment group in the ALPS Trial (RR 1.60 (95% CI 1.37-1.87)), also likely varies among population subgroups (e.g., gestational age).^10,14,15^ We could not investigate the absolute risks of hypoglycemia across gestational age and mode of delivery because these data are not publicly available due to ongoing data collection at the time of this analysis.^13^ Furthermore, there is no data available to inform individualized counseling on the associations between long-term neurocognitive or developmental risks and antenatal steroids in the late preterm period. Available evidence in other populations is mixed, though a single dose of antenatal steroids in the early preterm period has not been shown to increase long-term neurodevelopment risks.^16–21^

The study is strengthened by its use of rigorously collected data as part of a large randomized controlled trial, allowing us to estimate the rates of respiratory morbidity among those exposed and unexposed to antenatal steroids in the late preterm period. However, it is limited in that the trial enrollees may not reflect the general population at risk for late preterm birth or considering antenatal steroids. As mentioned, the baseline risks of respiratory morbidity are presented for individuals with a high likelihood of delivery in the late preterm period.^10^ The baseline risks may vary in a population without the same likelihood of preterm delivery. Similarly, the original trial was not powered to detect differences within population subgroups. As such, the number of patients within our strata varies, some of which are small, leading to considerable uncertainty. To help quantify this uncertainty, 95% confidence intervals are provided in Tables 1 and 2 and in the online calculator. Other factors may also contribute to a neonate’s risk for respiratory morbidity. However, this study focused on two patient-specific factors that would be known at the time of presentation and have previously been shown to have a significant effect on risk in the literature; future studies could try to improve the performance of the predictive model for respiratory morbidity using other factors, though the baseline challenge of predicting who and when a patient will deliver in the preterm period likely will continue to limit model discrimination. Last, we acknowledge that many non-patient factors may affect clinical decision-making surrounding ALPS (e.g., hospital resources, etc.); this information and the visualization tool are intended to help conceptualize the absolute risk reduction of respiratory morbidity with ALPS, which may inform one aspect of the shared decision-making.

In conclusion, the absolute risk of neonatal respiratory morbidity among patients at high risk for late preterm delivery varies considerably by gestational age at presentation and planned mode of delivery. Assuming no treatment effect heterogeneity (i.e., a constant relative risk reduction among all subgroups), patients with a low baseline risk (e.g., those presenting in the 36^th^ week and planning a vaginal delivery) can expect to obtain a considerably smaller absolute benefit from steroid administration compared to patients at the highest risk of neonatal respiratory morbidity (e.g., those presenting in the 34^th^ week and planning a cesarean delivery). ; Accurate understanding of these anticipated benefits and potential risks is critical to patients’ making decisions based on informed patient preferences. Studies adequately powered to compare the risks and benefits of antenatal late preterm steroids among population subgroups, and their effectiveness outside the context of a clinical trial, are needed to increase the precision of shared decision-making and public policy surrounding late preterm steroids.

## Data Availability

All data used in this study are publicly available via the NICHD's Data and Specimen Hub (https://dash.nichd.nih.gov/).

## Data Access, Responsibility, and Analysis

Dr. Clapp had full access to all the data in the study and takes responsibility for the integrity of the data and the accuracy of the data analysis.

## Data Sharing

Data used in this study is publicly available at https://dash.nichd.nih.gov/.

## Acknowledgments

The authors have no other acknowledgments.

