## Supplementary material for "Neonatal Respiratory Morbidity among Late Preterm Births by Planned Mode of Delivery and Gestational Age": Online Supplement

eTable 1: Sample sizes of strata

| Week of Gestation at Presentation | Planned Vaginal Delivery n=2,351 | Planned Cesarean Delivery n=474 |
|---|---|---|
| 34 weeks | 644 (27.4%) | 98 (20.8%) |
| 35 weeks | 887 (37.7%) | 213 (44.9%) |
| 36 weeks | 820 (34.9%) | 163 (34.3%) |

eTable 2: Distribution of patent factors between the control and treatment groups

| Factor | Control n=1,399 | Treatment n=1,426 | P value |
|---|---|---|---|
| Gestational age at randomization (days)[1] | 248.4 (5.5) | 248.6 (5.4) | 0.31 |
| Gestational age at randomization (weeks) | | | 0.21 |
| 34 weeks | 388 (27.7%) | 354 (24.8%) | |
| 35 weeks | 534 (38.2%) | 566 (26.7%) | |
| 36 weeks | 477 (34.1%) | 506 (35.5%) | |
| Planned Mode of Delivery | | | 0.70 |
| Vaginal | 1,173 (83.8%) | 1,178 (82.6%) | |
| Cesarean | 226 (16.2%) | 248 (17.4%) | |

[1] Presented as mean (standard deviation).

eTable 3: Observed rates and corresponding 95% confidence intervals of respiratory morbidity among the neonates not exposed and exposed to antenatal steroids, stratified by planned mode of delivery and week of gestation

| Week of Gestation at Presentation | Planned Vaginal Delivery | | Planned Cesarean Delivery | |
|---|---|---|---|---|
| | No ALPS % (95% CI) | ALPS % (95% CI) | No ALPS % (95% CI) | ALPS % (95% CI) |
| 34 weeks | 20.5 (16.2-24.8) | 17.6 (13.3-21.9) | 39.2 (25.6-52.7) | 31.9 (18.4-45.4) |
| 35 weeks | 12.1 (9.1-15.2) | 10.2 (7.4-13.0) | 25.8 (17.0-34.5) | 12.7 (6.1-18.0) |
| 36 weeks | 5.3 (3.1-7.5) | 6.2 (3.9-8.5) | 16.7 (8.3-25.0) | 11.8 (4.9-18.7) |

ALPS, antenatal late preterm steroids.

eTable 4: Unadjusted and adjusted risk of respiratory morbidity

| Model Term | Unadjusted Risk Ratio (95% CI) | Adjusted Risk Ratio (95% CI) |
|---|---|---|
| Treatment Group | | |
| Placebo | Reference | Reference |
| Antenatal Steroids | 0.81 (0.66, 0.98) | 0.81 (0.68, 0.97) |
| Planned Mode of Delivery | | |
| Vaginal | Reference | Reference |
| Cesarean | 1.79 (1.45, 2.20) | 1.90 (1.55, 2.33) |
| Week of Gestation (days) | 0.92 (0.91, 0.94) | 0.90 (0.90, 0.94) |

CI, confidence interval.

eTable 5: Observed rates and corresponding 95% confidence intervals of severe respiratory complications among the neonates not exposed to antenatal steroids, stratified by planned mode of delivery and week of gestation

| Week of Gestation at Presentation | Planned Vaginal Delivery | | Planned Cesarean Delivery | |
|---|---|---|---|---|
| | No ALPS | ALPS | No ALPS | ALPS |
| 34 weeks | 15.7 (11.8-19.6) | 13.7 (9.8-17.5) | 37.3 (23.9-50.7) | 23.4 (11.1-35.6) |
| 35 weeks | 10.5 (7.6-13.4) | 6.2 (4.0-8.5) | 23.7 (15.2-32.2) | 9.5 (4.1-14.8) |
| 36 weeks | 4.3 (2.3-6.2) | 3.3 (1.6-5.0) | 12.8 (5.4-20.3) | 8.2 (2.4-14.1) |

ALPS, antenatal late preterm steroids.

eTable 6: Unadjusted and adjusted risk of severe respiratory complications

| Model Term | Unadjusted Risk Ratio (95% CI) | Adjusted Risk Ratio (95% CI) |
|---|---|---|
| Treatment Group | | |
| Placebo | Reference | Reference |
| Antenatal Steroids | 0.66 (0.53, 0.83) | 0.66 (0.53, 0.83) |
| Planned Mode of Delivery | | |
| Vaginal | Reference | Reference |
| Cesarean | 2.00 (1.58, 2.54) | 2.16 (1.71, 2.72) |
| Week of Gestation (days) | 0.92 (0.90, 0.94) | 0.92 (0.90, 0.94) |

CI, confidence interval.

eFigure 1: Cross-validated receiver operating characteristic curves for model predicting the primary outcome (respiratory morbidity)

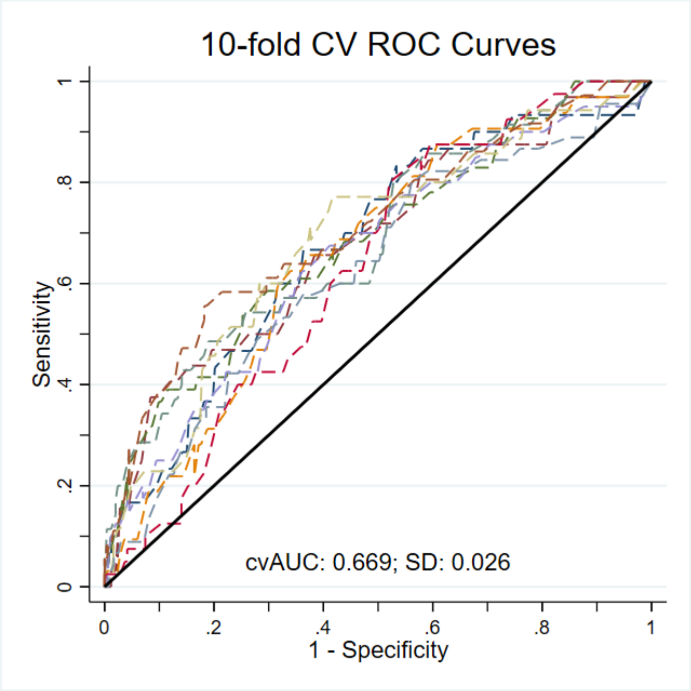


eFigure 2: Calibration plot for the model predicting the primary outcome (respiratory morbidity)

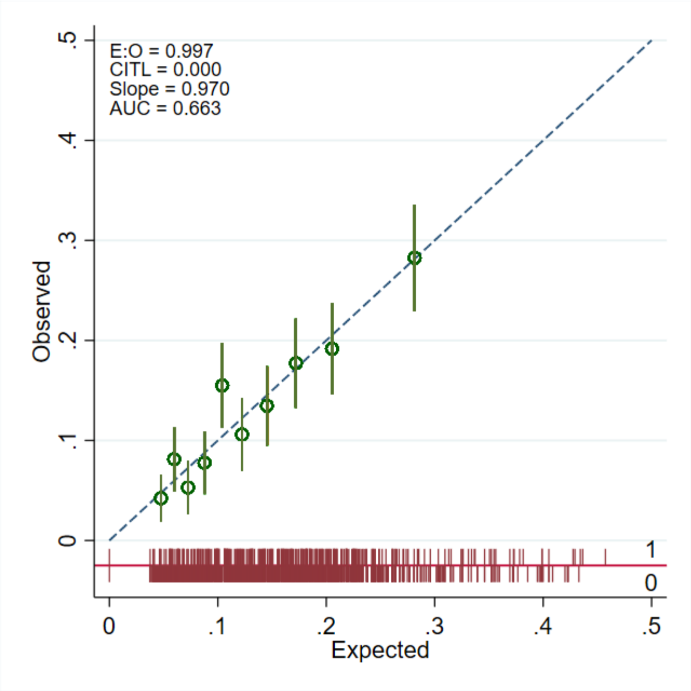

eFigure 3: Cross-validated receiver operating characteristic curves for the model predicting the secondary outcome (severe respiratory complications)

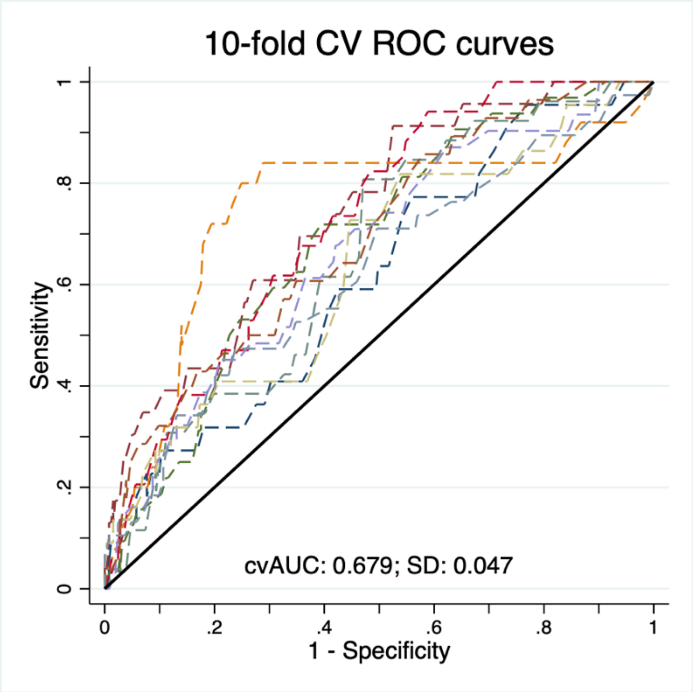


eFigure 4: Calibration plot for the model predicting the secondary outcome (severe respiratory complications)

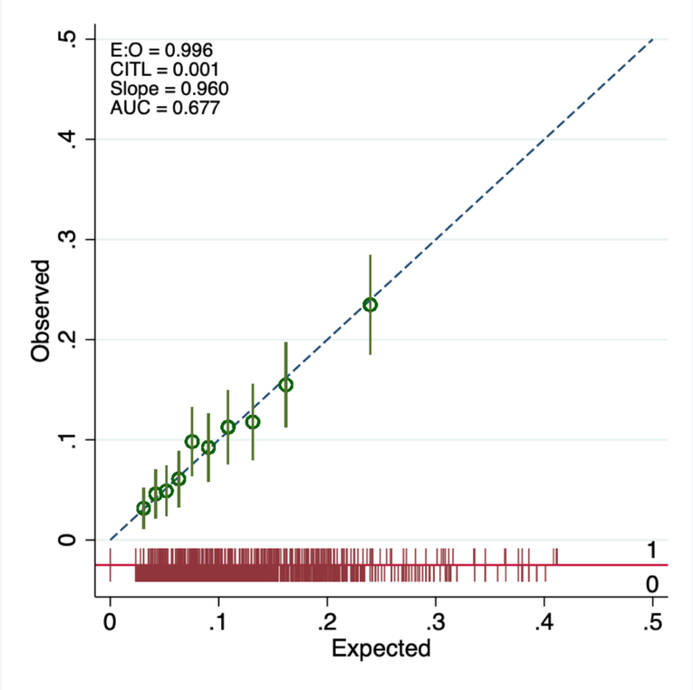
